# Relationship-centred care for people living with dementia in care homes. A rapid evidence summary

**DOI:** 10.1101/2024.04.15.24305839

**Authors:** Elizabeth Gillen, Deborah Edwards, Seren Roberts, Nia Davies, Isobel Davies, Jane Harden

## Abstract

Dementia is a progressive degenerative disease, typically affecting older adults for which there is currently no cure. Dementia is characterised by progressive impairment to several cognitive functions including memory and orientation, practical abilities and mood changes, all of which can impact personality and social relationships. The theory of social death has been explored for people living with dementia as the ability to maintain social interactions are threatened leading to a loss of social identity and exclusion and withdrawal from the wider community.

A relationship-centred care approach has been recommended to improve care for older people in long-term care, aiming to create environments conducive to relationships, considering the needs of the person living with dementia and also the staff and family members involved in their care. The ‘Senses Framework’ by Nolan was designed to promote relationships, acknowledging the experiences of all parties across six senses: security, continuity, belonging, purpose, fulfilment and significance. Utilising the Senses Framework has the potential to sustain meaningful relationships by fostering a sense of value and empowerment.

This Rapid Evidence Summary aims to explore the evidence assessing the benefits of a relationship triad approach (Senses Framework and other relationship-centred care approaches) in long term care settings (including care homes) for people living with dementia. Nine primary studies and one systematic review were identified.

The benefits of using a relationship-centred approach were mapped under each of the senses described within the Senses Framework, detailed separately for people living with dementia, relatives and care home staff. For people living with dementia, interventions using touch and music increased the sense of security and belonging reducing neuropsychiatric conditions such as agitation and aggression. Memory boxes were used to support a sense of significance and continuity and significant moments from the past brought pleasure and enabled meaningful conversations. For relatives and staff, improved communication and relationships increased confidence and trust and by working together they found that they could exert more influence and could develop into a powerful force for change.

## Wales Centre For Evidenced Based Care (WCEBC) Rapid Evidence Summary

### Relationship-centred care for people living with dementia in care homes

#### EXECUTIVE SUMMARY

##### What is a Rapid Evidence Summary?

This Rapid Evidence Summary is designed to provide an interim evidence briefing to inform further work and provide early access to key findings. It is based on a limited search of key resources and no quality appraisal or evidence synthesis was conducted, and the summary should be interpreted with caution.

##### Who is this summary for?

This Rapid Evidence Summary was conducted on behalf of Padraig McNamara, Mental Health and Vulnerable Groups, Welsh Government.

##### Background / Aim of Rapid Evidence Summary

Over 55 million people are reported to be currently living with dementia, with nearly 10 million new cases each year. This figure is expected to continue to rise significantly presenting one of the greatest challenges facing health and social care globally. A relationship-centred approach has been recommended to improve care for older people in long-term care homes. This approach advocates the creation of an environment conducive to relationships, that considers the needs of the person living with dementia together with the needs of staff and family members. This rapid evidence summary aims to explore the evidence assessing the benefits of a relationship triad approach (Senses Framework and other relationship-centred care approaches) in long term care settings (including care homes) for people living with dementia.

##### Key Findings

###### Recency of the evidence base

Searches were conducted in three databases (Medline, PsycINFO and CINAHL) from 2006 to September 2023.

###### Extent of the evidence base

▪ The searches conducted found **10** relevant studies:
  - Nine primary studies: qualitative (n=2), quantitative (n=3), mixed methods (n=3) and theoretical/conceptual study (n=1).
  - One secondary evidence: systematic review (n=1).

###### Key characteristics of the evidence base

▪ The primary studies were conducted in the UK (n=4), USA (n=3) Australia (n=1) and Belgium (n=1).
▪ The systematic review included six studies conducted in the USA (n=3), UK (n=1) Australia (n=1) and Sweden (n=1).
▪ The included studies were published between 2005 and 2023, with the majority (n=8) within the last 10 years.
▪ Study participants were a mixture of residents (n=8), care staff (n=7) and relatives (n=2).
▪ All studies investigated some aspect of relationship-based care with six focusing on the Senses Framework.

###### Benefits for people living with dementia

- An increased sense of **security** from interventions using touch.
- **Continuity** of support worker and avoiding injuries helped PLWD to **trust and feel safe** and **secure** within the care home environment.
- **Memory boxes**, **reminiscing** about past events and **individualised activities** gave PLWD a sense of **significance and continuity**, allowing them to feel known as **individuals** and hold **meaningful** conversations.
- Being **empowered** to make decisions and participate actively in their own care helped to preserve their **‘sense of self’** maintaining **independence and autonomy**.

###### Benefits for significant carers / relatives

- **Involvement** introduced a sense of **belonging and purpose** whereby the relative felt valued and respected and that their contribution could influence decision making.
- Involvement and **better relationships** with care home staff also increased relative’s **confidence** in the home and helped them **come to terms with their feelings of guilt** having placed a relative into long term care.
- Relatives coming together and communicating helped them see that they can **learn from each other**.
- **Continuity** of staff allowed relatives to feel able to **approach** staff to discuss health care needs. I**mproved dialogue** created a **better understanding** between them and provided opportunities for relatives to **support staff** in their caring role.

###### Benefits for care home staff

- **Recognition** from outside agencies helps to establish the **reputation** of the care home, helping to secure the home’s future.
- **Access** to managers, regular meetings with minutes and appraisals provided the opportunity to **air concerns** and receive **positive feedback**.
- **Understanding** and **managing risks** when providing care and feeling supported in their decision-making provided staff with a **sense of security**.
- Memory boxes and **personal information** helped staff to develop a **greater understanding** of PLWD and supported them in initiating **meaningful conversations**. Meaningful conversations also promoted a sense of purpose for staff.
- Staff and relatives found that **working together** increased their **sense of belonging** and helped them to develop a **powerful force for change** that could exert more influence.

##### Implications for practice

- The **Senses are more likely to be achieved** when they are applied in the context of a **relationship-centred approach to care**, rather than a person-centred model. This will require a change in practitioner thinking which see person-centredness as the foundation of care.
- The creation of an ‘**enriched’** environment of learning and care, as defined by the Senses Framework, has the potential to **‘transform’** healthcare professional’s views of what constitutes nursing and care, especially in relation to older people.
- The factors needed to create the Senses require ‘enriched care’ environments requires a range of **disciplines working collegiately** and with mutual respect. Current care environments need to **dismantle traditional hierarchies.**
- Most of the empirical work with the Senses Framework has been **completed with nursing staff**; however, it is potentially of **relevance across disciplines** and care settings.
- The Senses and relationship-centred care can provide a **framework for education** and practice to ensure the creation of ‘enriched’ environments of care in which the needs of **all groups** are accorded **equal value**, status and significance.

## 1. CONTEXT / BACKGROUND

Over 55 million people are reported to be currently living with dementia, with nearly 10 million new cases each year; it is the seventh leading cause of death globally (World Health Organisation 2023). This number is expected to increase significantly with numbers almost doubling every 20 years to reach 78 million in 2030 and 139 million in 2050 (Alzheimer’s Disease International 2023b) signifying one of the greatest current challenges facing health and social care globally (Woods et al. 2023).

Dementia is an umbrella term for neurodegenerative brain disorders typically affecting older adults and characterized by cognitive and behavioural symptoms (Woods et al. 2023). Alzheimer’s disease and vascular dementia (and mixed-type) are the most common forms of dementia. Dementia is a progressive degenerative disease affecting day-to-day life abilities. Progressive impairment in several cognitive functions is observed in people living with dementia, including memory and orientation, language skills reasoning, judgement, visuo-spatial skills, executive functioning, and practical abilities; personality and social relationships may also be affected (Woods et al. 2023), along with mood changes (Alzheimer’s Disease International 2023a). Despite some pharmacological therapies (such as acetylcholinesterase inhibitor medications) being reported to offer symptom relief (Alzheimer’s Research UK 2022) and slow progression of dementia (Yiannopoulou and Sokratis 2012) there is currently no cure for dementia nor effective therapies to halt the progression of the disease (World Health Organisation 2023). Thus, non-pharmacological interventions and support are considered essential for enhancing well-being of those living with dementia (Woods et al. 2023).

The theory of social death has been explored with people living with dementia and their families (Nimmons et al. 2023). Three underlying notions of social death include: a loss of social identity, a loss of social connectedness and losses associated with disintegration of the body (Kralova 2015). Accordingly, social roles change and people’s ability to maintain their previous social interactions are threatened by their chronic condition as their body’s decline advances; and losing social identity leads some to be excluded or withdraw from the wider community. Applying this theory to experiences of those living with dementia and their carers, Nimmons et al. (2023) in their qualitative study found this theory no longer fits with the way dementia is viewed or understood, with greater emphasis now on positive views of living well with dementia instead of focussing on loss and social exclusion. They argue that social connectedness and social identity, as previously described in the form of personhood and citizenship, can be maintained when people living with dementia are seen and treated as socially active. Maintaining social identity and connectedness as part of living well with dementia should be a core focus of any care approach.

A relationship-centred care approach has been recommended to improve care for older people in long-term care (Nolan et al. 2006). The aim of relationship-centred care is to create an environment that is conducive to relationships and considers not only the needs of the person living with dementia (PLWD), but also the needs of the staff and family members involved their care. The ‘Senses Framework was designed to promote all-round relationships between patients (individuals) and those who support and care for them by acknowledging the experience of all parties across six senses: sense of security, continuity, belonging, purpose, fulfilment, and significance (Nolan 1997; Nolan et al. 2006). Utilising the Senses Framework’ in dementia care has the potential to enhance the overall care experienced by fostering a sense of value and empowerment among staff and family members, enabling them to sustain meaningful relationships with individuals living with dementia.

This is in stark contrast to the more commonplace approaches to dementia care where a more ‘transactional’ model of care is to be found (Salisbury 2020). The result may be a well-kempt individual but the lack of human interaction and a sense of belongingness have profound effects; both on the person living with dementia and the care worker (Nolan et al. 2008). This has a knock-on effect in that staff turn-over is high and forming meaningful therapeutic relationships is not possible, thus increasing the burden on the family (Nwadiugwu 2021).

To summarise, the fundamental premise of Nolan et al.’s vision of relationship-centred care is that good care can only be delivered when the ‘senses’ are experienced by all three groups involved: the person, their significant other and the care worker. This rapid evidence summary therefore aims to explore the evidence assessing the benefits of a relationship triad approach (Senses Framework and other relationship-centred care approaches) in long term care settings (including care homes) for people living with dementia. A relationship triad approach for the context of this review includes the person living with dementia, their significant carer and the care home staff member.

## 2. RESEARCH QUESTION(S)

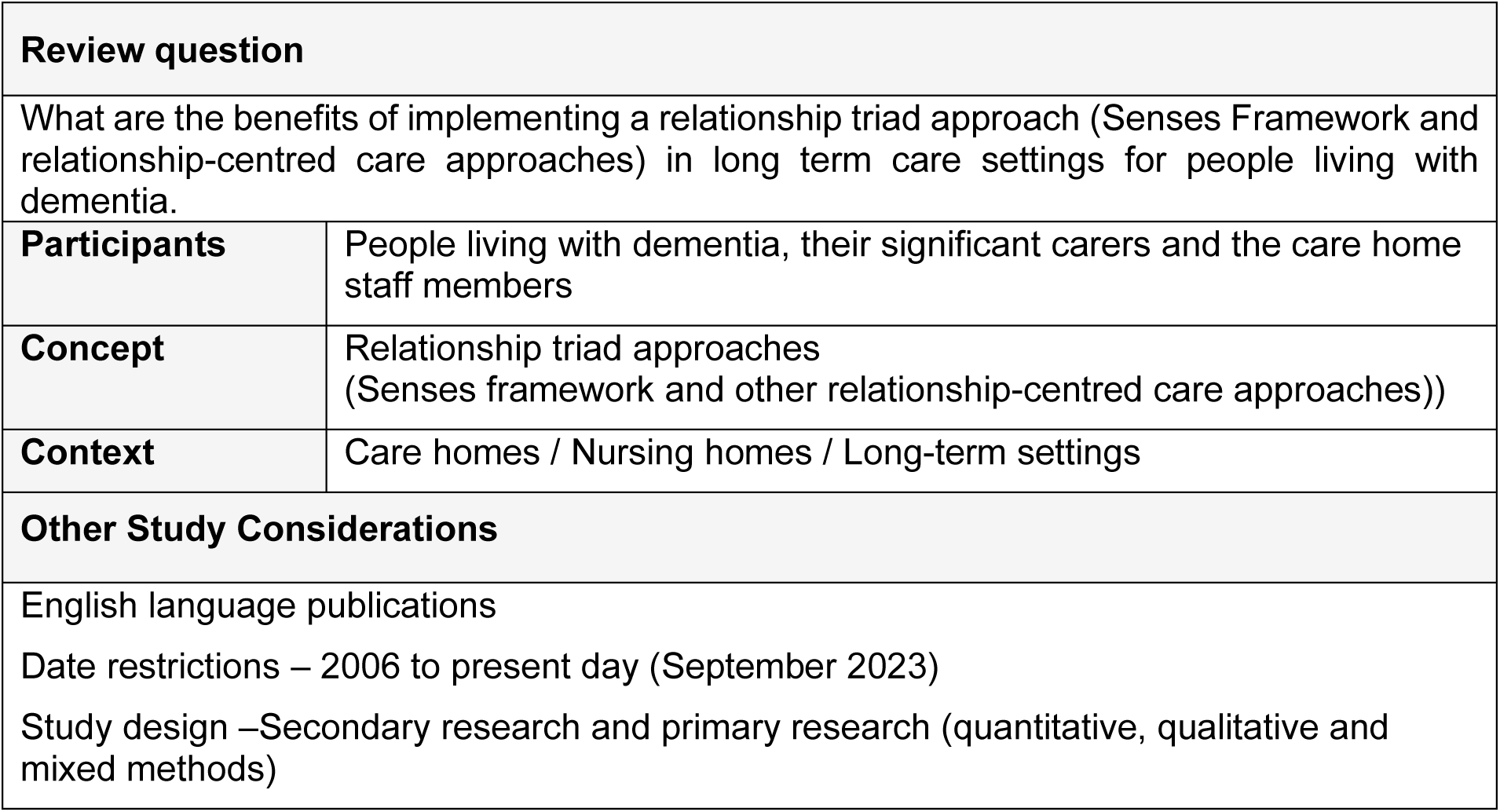

## 3. SUMMARY OF THE EVIDENCE BASE

### 3.1. Type and amount of evidence available

The evidence retrieved included nine primary studies and one systematic review. These included two qualitative studies (Allison et al. 2019,; Watson 2019), three quantitative studies (Gillis et al. 2019; Jablonski-Jaudon et al. 2016; Trinca et al 2021), three mixed methods studies (Aveyard & Davies. 2005; Brown Wilson et al. 2013; Ryan et al. 2008), one qualitative systematic review (Novy et al. 2023) and one theoretical/conceptual study (Taylor et al. 2014) investigated some aspect of relationship-centred care and of these six looked at the Senses Framework (Aveyard & Davies. 2005, Brown Wilson et al. 2013, Gillis et al. 2019, Ryan et al. 2008, Taylor et al. 2014, and Watson. 2019). A more detailed summary of included evidence can be found in Table 2 and a summary is provided below:

- A qualitative ethnographic study which was conducted in a nursing home **end-stage dementia** special care unit in the USA to investigate how the unit operated as a compassionate community using a **relationship-centred approach** for patients at the end-of-life (Allison et al. 2019).
- A qualitative descriptive study that evaluated the implementation of an action group in a nursing home in the North of England to investigate collaborative methods that could be used to improve the **experience of residents living with advanced dementia**, carers and staff within the home. The project was underpinned by the notion of relationship-centred care and the **Senses Framework** (Aveyard & Davies. 2005).
- A mixed methods study which was conducted across two care homes in the UK to produce, implement and assess a training programme aiming to enhance the **quality of care for dementia patients** based on relationship-centred care approaches and the **Senses Framework** (Brown Wilson et al. 2013).
- A qualitative study which was conducted across three nursing homes in Belgium that explored ways of collaborative working between residents, relatives, staff and researchers in order to create the most positive environment for people living with advanced dementia and their carers. The study evaluated **aggressive/agitated behaviours among dementia patients** and the use non-pharmaceutical interventions that focused on a person-centred team approach using the **Senses Framework** (Gillis et al. 2019).
- A quantitative descriptive study conducted in the USA to define a personalised method of providing **mouth care to people living with dementia,** focused on reducing threat perception and care resistant behaviours (Jablonski-Jaudon et al. 2016).
- A qualitative systematic review and meta-ethnographic study to understand the development of **relational based care** in long term care with a particular focus on residents living with **dementia** (Novy et al. 2023).
- A mixed methods study using questionnaires and interviews with **people living with dementia**, their carers and staff to evaluate the **respite service** provided by Community Health Sheffield using the **Senses Framework** (Ryan et al. 2008).
- Taylor et al. 2014 applied a conceptual framework of objectives aligned with the **Senses Framework** to two hypothetical scenarios (one of which was a living person with dementia in a care home) with various stakeholders involved **in mobility care** (including residents, care staff, physiotherapists, managers, and families) to develop mobility care practice improvement objectives.
- A cross-sectional multi-site Canadian study investigating associations between **relationship-centred care practices**, and the number of staff and residents at mealtimes in general and **dementia care** units in long-term care homes (Trinca et al. 2021).
- An qualitative ethnographic study in a social care home in Scotland to explore how care staff and **patients living with dementia** communicate and interact with each other and use theories to develop the **Senses Framework** and to advance relationship centred care in dementia at **end-of-life** (Watson. 2019).

### 3.2. Summary of the key findings

This section provides a narrative summary of the findings each of the included studies

- The study by Allison et al (2019) identified four themes that illustrated how a dementia special care unit operated as a companionate community for patients at the **end of life** using a **relationship-centred approach.** These were family as an ideal model for caregiving evoked by three relationship centred care techniques: reminiscence, verbal communication and non-verbal communication.
- Aveyard and Davies (2005) identified four themes from their study that was underpinned by the notion of **relationship centred care** and the **Senses Framework**. These were, creating a shared understanding, learning to value each other, becoming a powerful voice and moving forward. Implementing the **Senses Framework** was a useful guide for the development of collaborative care in long-term care of older people.
- A mixed method study by Brown Wilson et al (2013) assessed a training programme, based on the principles of **relationship-centred care** and the **Senses Framework** and identified five themes that included sense of security, belonging, purpose, a sense of significance and a sense of continuity. Staff members were able to gain a better understanding of the person with dementia by applying a biographical approach to care planning based on the principles of the **Senses Framework**
- There is a potential for developing individualised interventions based on the ABC method and the **Senses Framework** to address **agitated and aggressive behaviour** in residents living with dementia in nursing homes. The ABC method is a tool utilised by caregivers to understand residents needs and recognise challenging behaviours. (Gillis et al. 2019).
- Jablonski-Jaudon et al. 2016 successfully used a **relationship based care intervention** to deliver **oral hygiene** for older adults with dementia in nursing homes.. Mouth care procedures were carried out in a manner that brought enjoyment to the nursing home residents. was accomplished and completed in a manner enjoyable to nursing home residents. Relationships that were initially centred around the task of oral care, evolved into more intimate and responsive connections between the oral care practitioners and the nursing home residents.
- A qualitative systematic review by Novy et al (2023) looking at **relational care,** identified five transferable concepts. **Doing with versus doing for** whereby ‘doing with’ results in cooperative and synergetic care and promotes equality and empathy. This is broken down into staff responsiveness, being open, resident agency, inclusive communication and time.
- The **Senses Framework** was used as an analytical framework to represent the different perspectives of people living with dementia, their carers and staff that are necessary to consider to deliver **high quality respite care** (Ryan et al. 2008).
- The **Senses Framework** can be used to develop objectives of **mobility care practice** improvement and the authors highlighted the potential advantages of person-centred approaches in enhancing the resident-staff relationship (Taylor et al. 2013).
- In general care long term settings more staff was associated with improved **relationship-centred care during mealtimes**. however the same associations were not seen for dementia care units (Trinca et al 2021).
- The Senses Framework can support the provision of relationship-centred care in **palliative care for people living with dementia**. In the advanced stages of dementia, people are still able to fully experience interpersonal relationships with those who care for them (Watson. 2019).

### 3.3. Benefits of a relationship-centred care approach

This next section describes the benefits (highlighted within the included studies) of using a relationship-centred care approach mapped under each of the senses described within the senses framework (Nolan. 1997, Nolan et al, 2006). The benefits recognised under each of the senses have been detailed separately for people living with dementia, significant carers / relatives and care home staff

#### 2.3.1. A sense of security

– of feeling safe and receiving or delivering competent and sensitive care

*People living with dementia*:

- Interventions using **touch for a need for an increased sense of security** or music therapy for a need for an increased sense of belonging or purpose, can be effective in **reducing** the number and severity of neuropsychiatric symptoms including **agitation, aggression, depression and loss of decorum** (Gillis et al. 2019).
- For the resident (or PLWD), benefits might include a sense of security by **avoiding injury** and **feeling safe and comfortable** during care activity (Taylor et al. 2014).
- Where residents got to know the ‘face’ of the staff, it helped create a **sense of security and continuity** (Watson. 2019).

*Relatives:*

- Better relationships increased **relatives’ confidence and trust** in the home (Aveyard and Davies. 2005).
- The involvement in the action group had helped relatives **come to term with their feelings** about placing the older person into long-term care (Aveyard and Davies. 2005).
- Some evidence that involvement in the project might also **assist relatives at the end of the resident’s life** (Aveyard and Davies. 2005).
- Carers **gained a break** from caring and **felt confident with the quality of care** provided (providing safety and security) (Ryan et al. 2008).
- Secure in the knowledge that their relative is looked after and having a good time helps to **relieve any guilt felt** (Ryan et al. 2008).
- A sense of security by feeling comfortable that their loved one will receive best care (Taylor et al. 2014).

Staff:

- Outside interest in the home helps to **secure the home’s future** / **recognition** from outside agencies / helped to **establish a reputation** within the trust (Aveyard and Davies. 2005).
- Receiving positive feedback from relatives (Aveyard and Davies. 2005).
- **Openness and honesty** within meetings, with notes and minutes (Aveyard and Davies. 2005).
- A sense of security by **feeling safe and avoiding injury** when providing care, **understanding and managing risks**, and **feeling supported** to deliver best care (Taylor et al. 2014).

#### 2.3.2. A sense of continuity

– the recognition of biography, using the past to contextualise the present

*People living with dementia:*

- The **importance of reminiscing** about past events was stressed as an important way in which to attempt to evoke long-term memories that may still be present for care recipients (Allison et al. 2019).
- Memory boxes used to support a **sense of significance and continuity** for the person with dementia. The personal information helped staff to develop a **greater understanding**, giving the person with dementia **pleasure** (Brown Wilson. 2013).
- Individualised **activities to increase sense of continuity**, achievement and significance, were not effective in reducing neuropsychiatric symptoms but did **reduce agitation/aggression**. (Gillis et al. 2019).
- **Increased participation** in social life and **stimulation** that reflected personal preferences (Ryan et al. 2008).
- A sense of continuity allowed PWLD to feel that they were **known as individuals**. The continuity of support worker allowed the person with dementia to **get to know and trust them and to feel secure** (Ryan et al. 2008).
- The benefits of relational care for PLWD include being more visible in their care activities; being **empowered** to make decisions; **participating** in communication; and, having space to participate actively in their care by supporting them to try things for themselves, doing the things they can still do, interacting socially, and **preserving their sense of self** (Novy et al. 2023).
- A sense of continuity by being **comfortable and familiar** with staff providing care (Taylor et al. 2014).
- Embodied aspects of **selfhood**, such as wearing certain items of clothing were found to be important (Watson. 2019).
- When a resident who rarely speaks is spoken to, has their hand held and is included and as they reciprocate with a word of thanks or smile, something of that person’s remaining **embodied selfhood** and their agency ‘springs forth in joint activity’. They are being treated as an experienced body rather than a body object. Selfhood was seen as crucial to challenging social death and alleviating the distress this can cause in advanced dementia. (Watson. 2019).

*Relatives:*

- **Improved communication and enhanced relationships** between staff and relatives (Aveyard and Davies. 2005).
- Relatives coming together and communicating has helped them see that they can **learn from each other** (Aveyard and Davies. 2005).
- The **continuity** of support worker, use of biographical methods and support workers continued **efforts to engage** with the carer and person with dementia adds to the carer’s feelings of security and continuity (Ryan et al. 2008).
- The attention to **personhood** not only helps the person with dementia but **helps to improve the relative’s well-being** (Ryan et al. 2008).
- A sense of **continuity by knowing** the staff caring for their loved one and feeling **able to approach** care staff to discuss care needs (Taylor et al. 2014).

*Staff:*

- Memory boxes and personal information helped staff to develop a **greater understanding** of the person with dementia and supported them in **initiating and promoting more meaningful conversations** (Brown Wilson. 2013).
- Regular meetings of the action group with minutes. Provided **time to air concerns** and provide **positive feedback** to staff (Aveyard and Davies. 2005).
- Regular access to senior staff and managers (Aveyard and Davies. 2005).
- A sense of continuity by **getting to know the resident and their care needs** or by getting to know other staff who know the resident (Taylor et al. 2014).
- Knowing things about the person’s life were described as **‘handles to hold onto’** which helped them connect person-to-person, and to care (Watson. 2019).
- **Embodied knowing** was combined with **knowledge of the persons culture** in ways which helped caring run more smoothly (Watson. 2019).

#### 3.3.3. A sense of belonging

– opportunities to form meaningful relationships or feel part of a team

*People living with dementia:*

- The caring community demonstrated ingenuity in the creation of moments of connection with the men and women living on the SCU (verbal and non-verbal), revealing that ideal dementia care has a bidirectional focus in which members of the caring community display empathy and create **moments of mutual meaning** (Allison et al. 2019).
- Following the workshops there was an increase in staff responding that residents were ‘usually’ able to hold **meaningful conversations** with them (Brown Wilson. 2013).
- Significant moments from the past can bring pleasure to the present and allow for **meaningful conversations** (Brown Wilson. 2013).
- A sense of belonging by being acknowledged and feeling part of a team during care activities (Taylor et al. 2014).

*Relatives:*

- Relatives valued **having a place and a role** in the home (Aveyard and Davies. 2005).
- **Improved dialogue creates a better understanding** of staff problems and creates opportunities to provide support for staff (Aveyard and Davies. 2005).
- By spending more time together, staff and relatives felt they had **developed a greater understanding** of each other’s needs and consequently relationships improved (Aveyard and Davies. 2005).
- Carers/Relatives feel that they are an **essential part of the team**, ensuring a sense of belonging (Ryan et al, 2008).
- A sense of belonging by **feeling involved** in the resident’s care decision making (Taylor et al. 2014).
- For significant carer or family members, benefits may include **involvement in the care** provided for their loved ones by **increasing meaningful interactions**, supporting food intake and helping to reduce staff time constraints (Trinca et al. 2021).

*Staff:*

- Working together on fundraising initiatives (Aveyard and Davies. 2005).
- Relatives providing support for staff (Aveyard and Davies. 2005).
- Staff reported an improvement in working together as a team with more opportunities to discuss the care of residents (Brown Wilson. 2013).
- Meaningful conversations not only contributed to improved quality of care for the resident but may also promote a sense of purpose for staff (Brown Wilson. 2013).
- Benefits for mouth care providers were successfully performing mouth care in an enjoyable way, **developing relationships** with residents through continuity, problem solving on the spot (bite blocking) and honing their skills in threat reduction strategies (Jablonski-Jaudon et al. 2016).
- The benefits for staff relate to: **seeing the whole person** and recognising that the **PLWD is a key player in the care relationship**; developing skills of “(1) an openness to the resident during the care activities, (2) an ability to recognize when a resident is able to make their own decisions and to do things for themselves, and (3) a sensitivity to non-verbal communication as a means towards more inclusive communication”, without notable time demands; and, have a more “cheery and personal demeanour”, genuine interest and enjoyment, and encouraging recall through a ‘doing with’ approach with active inclusive communication, and responsive care (Novy et al. 2023).
- Cohesion and mutually supportive work environment (Ryan et al. 2008).
- A sense of belonging by being part of a team or being instrumental in delivering best care (Taylor et al. 2014).
- Body work: many care staff saw **body work** as an opportunity to **connect with the person** and foster a sense of belonging (Watson. 2019).

#### 3.3.4. A sense of purpose

– opportunities to engage in powerful activities or to have a clear set of goals to aspire to

*People living with dementia:*

- Benefits for PLWD included: **successfully engaging** with and receiving mouth care, **collaborating** with the mouth care provider, enjoying mouth care and developing relationships through continuity of mouth care provider (Jablonski-Jaudon et al. 2016).
- A sense of purpose by being able to **actively participate** in care activities with assistance for comfort and function and maintain as much **independence and autonomy** as possible (Taylor et al. 2014).
- Relationship-centred care during mealtimes may benefit PLWD by promoting mealtime **independence** and engaging residents in mealtime activities, thereby increase quality of life, wellbeing, and potentially food intake (Trinca et al. 2021).
- Experienced residents led to **embodied knowing** (learning about care needs) (Watson. 2019).

*Relatives:*

- **New purpose** in visiting the home (Aveyard and Davies. 2005).
- Able to **contribute** in a different way (Aveyard and Davies. 2005).
- Relatives have a sense of **being able to change** things (Aveyard and Davies. 2005).
- A sense of purpose by feeling that they **contribute to the care being provided**; a sense of **achievement** to assist the resident to retain function and independence (Taylor et al. 2014).

*Staff:*

- Specific development projects (Aveyard and Davies. 2005).
- Introduction of appraisal (Aveyard and Davies. 2005).
- The groups orientation to **promote change encouraged a shared sense of purpose** and responsibility (Aveyard and Davies. 2005).
- Staff and relatives found that by working together, they had **developed into a powerful force for change and could exert more influence** (Aveyard and Davies. 2005).
- The ‘**powerful voice’** contributed to the project’s ability to move things forward (Aveyard and Davies. 2005).
- Following the workshops, staff reported improvement in being **kept informed** and working without being hurried (Brown Wilson et al. 2013).
- A sense of purpose by **understanding and meeting the resident’s care needs** and contributing to safe care culture (Taylor et al. 2014).

#### 3.3.5. A sense of fulfilment

– achieving meaningful or valued goals and feeling satisfied with one’s efforts

*People living with dementia:*

- Instils a **sense of purpose and achievement** for the people living with dementia as well as their carers (Ryan et al. 2008).
- A sense of **achievement** by **maintaining function and independence** and recognising successful care adaptations as needed (Taylor et al. 2014).

*Relatives:*

- New challenges and opportunities for relatives resulting in **new achievements** (Aveyard and Davies. 2005).

*Staff:*

- Staff described how they had become **more aware of the value** placed on their work by relatives (Aveyard & Davies. 2005).
- The sense of purpose and achievement is linked to support workers believing that they are **making a difference** to both the person with dementia and their carer/relative (Ryan et al. 2008).
- A sense of achievement by **safely assisting residents to retain function and independence**, and to **support other staff** in their sense of security and to provide best care (Taylor et al. 2014).

#### 3.3.6. A sense of significance

– to feel that you matter, and that you are valued as a person

*People living with dementia:*

- Person with dementia involved in decision making (Ryan et al. 2008).
- A sense of significance by **feeling valued** including contributions to own care activities (Taylor et al. 2014).

*Relatives:*

- The work of the project provided a **sense of belonging and purpose** for relatives and that their **contribution was valued** (Aveyard and Davies. 2005)
- Relatives had a place and a role and could **influence decision making** (Aveyard and Davies. 2005).
- Staff show that they **value relative’s contributions** to care. Also, relatives had a better understanding of staff problems and created an opportunity to provide support (Aveyard and Davies. 2005).
- A sense of significance by **feeling valued and respected** by staff for providing support for the care their loved one receives (Taylor et al. 2014).

*Staff:*

- Everybody’s **views respected** and acted upon (Aveyard and Davies. 2005).
- Opportunities for **mutual appreciation** (Aveyard and Davies. 2005).
- **Recognition** from outside agencies (Aveyard and Davies. 2005).
- The action group meetings **provided time to air concerns** and provide positive feedback to staff (Aveyard and Davies. 2005).
- Staff reported an increase in their **work being valued** by both colleagues and families (Brown Wilson et al. 2013).
- Sense of significance contributes to both **job satisfaction and morale** but also to their sense of who they are, both as a team and individually (Ryan et al. 2008).
- A sense of significance by **feeling valued and respected by residents** for assisting them in their care, **feeling valued by other including supervisors and managers** for the care they provide and feeling respected for their knowledge of best care provision (Taylor et al. 2014).
- **Rewarding encounters** foster a sense of achievement for staff and a sense of significance and belonging for both the person with dementia and the staff caring for them (Watson. 2019).

## Data Availability

All data produced in the present study are available upon reasonable request to the authors

## Abbreviations

Acronym: Full Description
PLWD: People Living With Dementia

## 4. RAPID EVIDENCE SUMMARY METHODS

Three databases were searched: Medline (Ovid), PsycINFO (Ovid), CINAHL (EBSCO) in September 2023 and the search strategies are provided in Appendix I. Searches were limited to English language publications. Search hits were screened for relevance by two reviewers and exclusions noted (see Appendix 2). The flow of citations through the review are displayed Figure 1 in a PRISMA flow diagram (Page et al. 2021).

**Figure 1:**
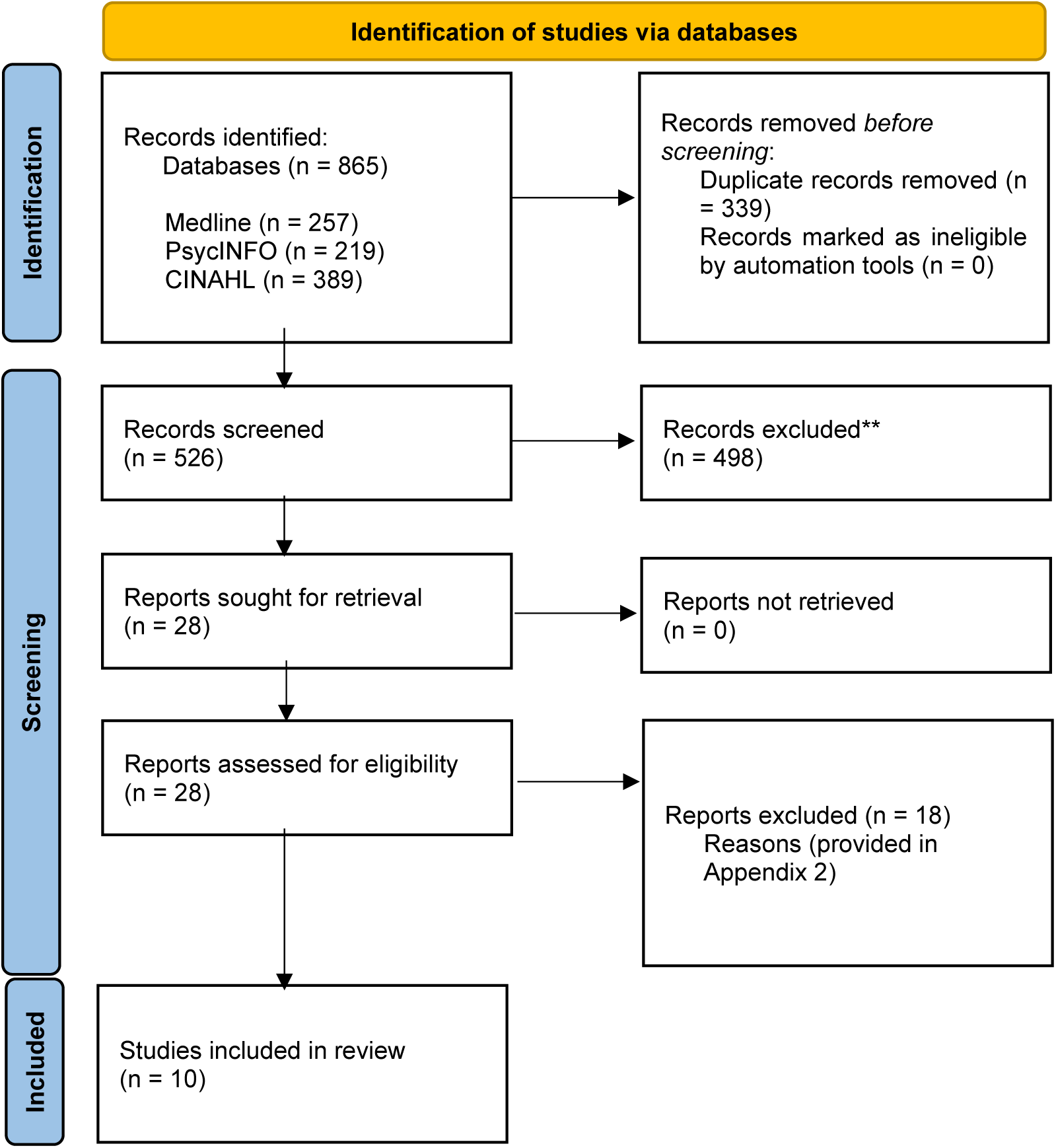
PRISMA flow diagram:

No formal quality assessment was conducted. Citation, recency, evidence type and key findings were tabulated for all relevant primary and secondary research identified in this process by one reviewer and checked for accuracy by a second.

**Table 1:** Summary of included primary research evidence.

| Citation | Study design / aims / geographical location | Methods / setting |
| --- | --- | --- |
| Allison et al. 2019<br>Caring community and relationship centred care on an end-stage dementia special care unit<br>https://dx.doi.org/10.1093/ageing/afz030 | <u>Study design</u><br>Qualitative study (ethnographic)<br><u>Aims</u><br>To investigate how a nursing home end-stage dementia special care unit operated as a compassionate community for patients who were at the end of life<br><u>Geographical location</u><br>USA | <u>Participants</u><br>Residents (n=19)<br>Care staff (n=23)<br>Volunteers (n=2)<br>Family members (n=3)<br><u>Methods</u><br>Ethnographic case study methodology was used to investigate aspects of care within the nursing home.<br>>175 hours of scheduled activity observations<br>30 hours of caregiving observations<br>Interviews<br><u>Setting</u><br>Nursing home end-stage dementia special care unit |
| Aveyard and Davies 2006<br>Moving forward together: evaluation of an action group involving staff and relatives within a nursing home for older people with dementia<br>https://doi.org/10.1111/j.1748-3743.2006.00005.x | <u>Study design</u><br>Mixed methods study<br><u>Aims</u><br>To explore ways of collaborative working between residents, relatives, staff and researchers in order to create the most positive environment for people with advanced dementia and their carers<br><u>Geographic Location</u><br>North of England, UK | <u>Participants</u><br>Care staff (n=18)<br>Relatives (n=7)<br><u>Methods</u><br>The project was underpinned by the notion of relationship-centred care and the Senses Framework<br>Semi structured interviews<br><u>Setting</u><br>Specialist nursing home for older people with dementia |
| Brown Wilson et al. 2013<br>The senses in practice: enhancing the quality of care for residents with dementia in care homes<br>https://doi.org/10.1111/j.1365-2648.2012.05992.x | <u>Study design</u><br>Mixed methods study<br><u>Aims</u><br>To develop, deliver, and evaluate a training programme based on the Senses Framework that | <u>Participants</u><br>Care staff (n=11) from four units within the care home<br><u>Methods</u><br>Eight workshops focusing on the principles on relationship centred care |
|  | encouraged staff to develop dementia care practice using the principles of relationship-centred care<br>The evaluation of the training programme intended to capture the perspectives of staff, residents with dementia, and relatives<br><u>Geographical location</u><br>North West of England, UK | Pre- and post evaluation involving the perspectives of residents with dementia, their families and staff was planned with CARE Profiles being used prior to the workshops starting and three months after staff had completed the workshops<br>A focus group of staff were recruited to assess the practice/training context using case examples after the study<br><u>Setting</u><br>One care homes |
| Gillis et al. 2019<br>A person-centred team approach targeting agitated and aggressive behaviour amongst nursing home residents with dementia using the Senses Framework<br>https://doi.org/10.1111/opn.12269 | <u>Study design</u><br>Quantitative study (pre-test / post-test))<br><u>Aims</u><br>To test a person-centred team approach for addressing agitated or aggressive behaviour amongst nursing home residents with dementia<br><u>Geographical location</u><br>Belgium | <u>Participants</u><br>Residents diagnosed with dementia who expressed agitated or aggressive behaviour (n=65)<br><u>Methods</u><br>Residents were allocated one of the three interventions, based on one of the six needs in the Senses Framework and the ABC method (the ABC method is a tool utilised by caregivers to understand residents needs and recognise challenging behaviours)<br>Residents with a need for sense of security were given therapeutic touch<br>Residents with an increased need for a sense of belonging or a sense of purpose were given group music sessions<br>Residents with a need for more sense for continuity, fulfilment or significance were given an individualised meaningful activity<br>Neuropsychiatric Inventory-Nursing Home version (NPI-NH) – measures and rates behavioural and psychological symptoms of dementia<br>The Cohen-Mansfield Agitation Inventory (CMAI) – measures the prevalence of agitation in the behaviour of the residents<br><u>Setting</u><br>Three nursing homes in six different wards |
| Jablonski-Jaudon et al. 2016 | <u>Study design</u> | <u>Participants</u> |
| Maturation of the MOUTH Intervention: from reducing threat to relationship-centered care<br>https://doi.org/10.3928/00989134-20160212-05 | Quantitative study (descriptive)<br><u>Aims</u><br>To describe a personalized practice originally conceived as a way to prevent and minimise care resistant behaviours to provide mouth care to older adults with dementia<br><u>Geographical location</u><br>USA | Residents with a diagnosis of dementia (n=46)<br>Mouth care providers (undergraduate students and nursing assistants who were hired specifically for the study)<br>2,275 mouth care sessions<br><u>Methods</u><br>Participants were randomised into either an experimental group or an active control group and both received twice-daily mouth care for 3 weeks<br>Control group received oral care according to the a standardised protocol<br>Experimental group received the full MOUTH intervention<br>For three weeks, research staff provided twice-daily oral care to both groups<br><u>Setting</u><br>Nursing homes (n=9) |
| Novy et al. 2021<br>Relational care, dementia and communication challenges in long-term care: a meta ethnography<br>https://doi.org/10.1017/S0714980822000253 | <u>Study design</u><br>Qualitative systematic review (5 studies)<br><u>Aims</u><br>To synthesis qualitative studies in order to understand the development of relational based care in long term care and in particular the participation of residents with dementia.<br><u>Geographical location</u><br>Australia (n=1), USA (n=3), UK (n=1), Sweden (n=1) | <u>Participants</u><br>People living with dementia in a residential care setting.<br><u>Methods</u><br>Used meta-ethnography to synthesise qualitative research. Conducted a comprehensive search of MEDLINE, CINAHL, PSYCinfo and Embase<br><u>Setting</u><br>Residential care |
| Ryan et al. 2008<br>Using the senses framework to achieve relationship-centred dementia care services: A case example<br>https://doi.org/10.1177/1471301207085368 | <u>Study design</u><br>Mixed methods study<br><u>Aims</u><br>To describe the development of a new service for people with dementia and their carers in a large post-industrial city in the north of England, UK | <u>Participants</u><br>People living with dementia, their carers and service providers (n=ns)<br><u>Methods</u><br>Questionnaires and interviews with<br><u>Setting</u> |
|  | <u>Geographical location</u><br>Sheffield, UK | Community (Community Health Sheffield) |
| Taylor M<br>Geron et al. 2013<br>Quality Improvements in resident mobility care: using person- and relationship-centered frameworks<br>https://doi.org/10.1093/geront/gnt082 | <u>Study design</u><br>Theoretical/conceptual study<br><u>Aims</u><br>To discover an appropriate person- or relationship-centred framework to improve mobility care in nursing homes.<br><u>Geographical location</u><br>Australia | <u>Participants</u><br>Nursing home residents (and those involved in mobility care practices within nursing homes)<br><u>Methods</u><br>Review and analysis of research literature and guidelines underpinned by Senses Framework and applied to two scenarios to discuss the challenges in mobility care and the application of person- and relationship-centred approaches in mobility care using the Senses Framework.<br><u>Setting</u><br>Nursing homes |
| Trinca et al. 2021<br>Making the Most of Mealtimes (M3): association between relationship-centred care practices, and number of staff and residents at mealtimes in Canadian long-term care homes<br>https://doi.org/10.1016/j.jamda.2020.11.020 | <u>Study design</u><br>Quantitative study (descriptive)<br><u>Aims</u><br>(1) to determine number of staff or residents associated with relationship-centred care practices at mealtimes<br>(2) to assess whether the association between number of staff and relationship-centred care is moderated by number of residents and family/volunteers, profit status or chain affiliation<br><u>Geographical location</u><br>Alberta, Manitoba, New Brunswick, and Ontario | <u>Participants</u><br>Residents (n=639) on general and dementia care units<br><u>Methods</u><br>Secondary data analysis of M3 study data in which residents from each home were randomly selected and recruited.<br>- MealTime Scan (MTS) – number of staff (i.e., plating / delivering meals or providing mealtime assistance), residents, and family/volunteers in the dining room<br>- 17-items on the Mealtime Relational Care Checklist (M-RCC) which is part of the MTS which provides information mealtime interactions<br>- 7-point Cognitive Performance Scale - resident cognition<br>10-item Edinburgh Feeding Evaluation in Dementia Scale – Level of physical eating assistance required<br><u>Setting</u><br>General (n=58) and dementia care units (n=24) from 32 care homes |
| Watson 2019<br>Developing the senses framework to support relationship-centred care for people with advanced dementia until the end of life in care homes<br>https://doi.org/10.1177/1471301216682880 | <u>Study design</u><br>Qualitative study (ethnographic)<br><u>Aims</u><br>To examine, through the lenses of embodied and interembodied selfhood, the ways care staff and people with dementia living in a care home relate to each other and how these theories can develop the Senses Framework and promote high quality relationship-centred care with people with dementia until the end-of-life<br><u>Geographical location</u><br>Scotland, UK | <u>Participants</u><br>Residents with advanced dementia at end-of-life (n=20)<br>Care staff (n=33)<br><u>Methods</u><br>Ethnographic approach<br>Observations (195 hours)<br>Interviews with care staff (n=24)<br>Group discussions with care staff (n=3)<br>Examination of care plans and daily records kept on residents participating in focussed observation<br><u>Setting</u><br>Specialist dementia social care home (n=1) |

## 5. ABOUT THE WALES CENTRE FOR EVIDENCE BASED CARE (WCEBC)

The WCEBC promotes evidence-based practice through the development and evaluation of internationally excellent systems for evidence appraisal, translation and utilisation.

We operate with a core team in the School of Healthcare Sciences, Cardiff University, and are led by [Dr Deborah Edwards](https://www.cardiff.ac.uk/people/view/39125-edwards-deborah) and [Dr Clare Bennett](https://www.cardiff.ac.uk/people/view/1131115-Bennett-Clare).

We work closely with the following organisations: JBI, European Cancer Organisation, World Health Organisation Collaborating Centre for Midwifery Development and the Wales COVID-19 Evidence Centre.

**Co-Directors:**

Dr Deborah Edwards Dr Clare Bennett

**Contact Email:**

**Website:** https://www.cardiff.ac.uk/research/explore/research-units/wales-centre-for-evidence-based-care

## 6. APPENDICES

## Appendix 1: Search strategies

**Medline (Ovid) 08.09.2023**

| # | Query | Results |
| --- | --- | --- |
| 1 | *Dementia/ | 51,019 |
| 2 | *Alzheimer Disease/ | 98,827 |
| 3 | (dementia or alzheimer*).tw. | 271,712 |
| 4 | 1 or 2 or 3 | 282,510 |
| 5 | (relation* adj2 (care or approach*)).tw. | 5,758 |
| 6 | senses framework.tw. | 25 |
| 7 | triad care.tw. | 2 |
| 8 | 5 or 6 or 7 | 5,777 |
| 9 | 4 and 8 | 291 |
| 10 | limit 9 to (english language and yr="2006 -Current") | **257** |

**PsycINFO (Ovid) 08.09.2023**

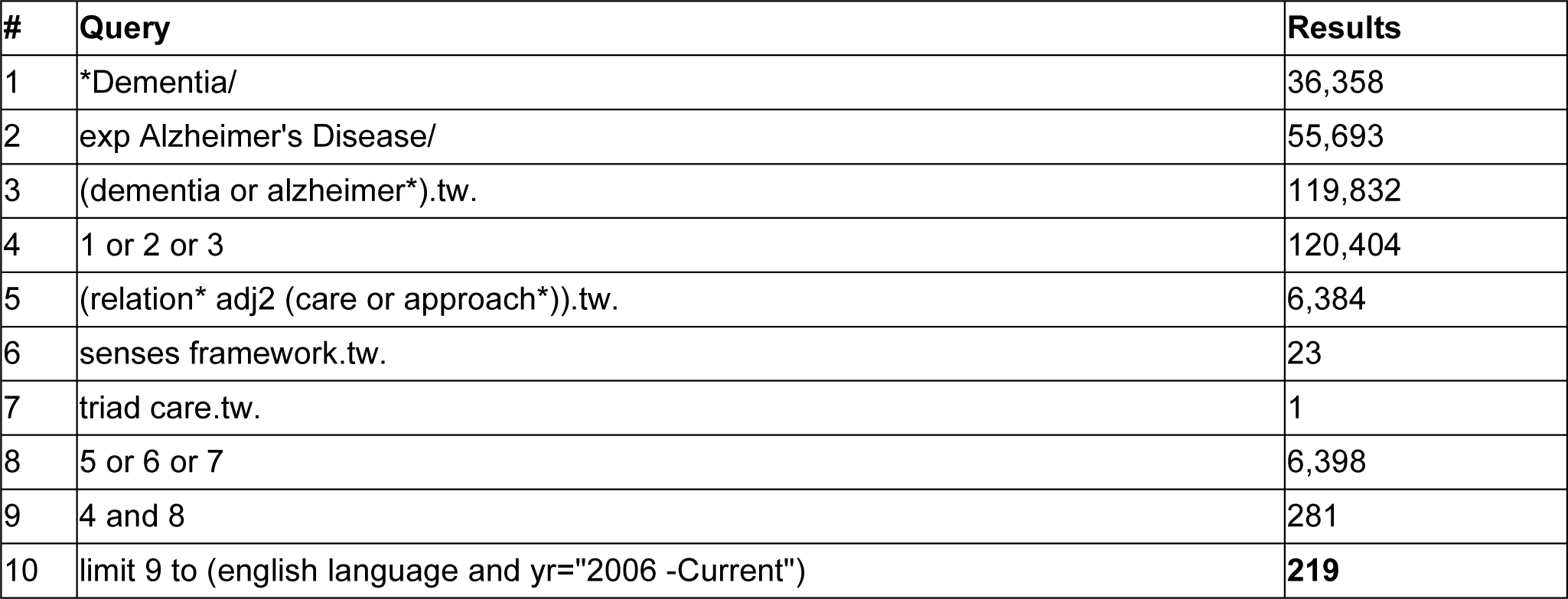

| # | Query | Results |
| --- | --- | --- |
| 1 | *Dementia/ | 36,358 |
| 2 | exp Alzheimer's Disease/ | 55,693 |
| 3 | (dementia or alzheimer*).tw. | 119,832 |
| 4 | 1 or 2 or 3 | 120,404 |
| 5 | (relation* adj2 (care or approach*)).tw. | 6,384 |
| 6 | senses framework.tw. | 23 |
| 7 | triad care.tw. | 1 |
| 8 | 5 or 6 or 7 | 6,398 |
| 9 | 4 and 8 | 281 |
| 10 | limit 9 to (english language and yr="2006 -Current") | **219** |

**CINAHL (EBSCO) 08.09.2023**

| # | Query | Results |
| --- | --- | --- |
| 1 | (MM "Dementia") | 33,500 |
| 2 | (MM "Alzheimer's Disease") | 28,826 |
| 3 | TI (dementia or alzheimer*) OR AB (dementia or alzheimer*) | 93,444 |
| 4 | 1 or 2 or 3 | 99,994 |
| 5 | TI ((relation*) N2 (care or approach*)) OR AB ((relation*) N2 (care or approach*)) | 8,832 |
| 6 | TI "senses framework" OR AB "senses framework" | 28 |
| 7 | TI "triad care" OR AB "triad care" | 2 |
| 8 | 5 or 6 or 7 | 8,851 |
| 9 | 4 and 8 limited to (english language and yr="2006 -Current") | **389** |

## Appendix 2: Studies excluded at full-text screening

1. Bradley et al. 2015. Crossing the bridge: connecting with people in the later stages of dementia. Reason for exclusion: opinion article which talks about relationships but doesn’t directly talk about or use the principles of relationship based care and the Senses Framework.
2. Coppalle et al. 2018. Helping caregivers of people with dementia: a need to renew theoretical frameworks in France. *Reason for exclusion:* English abstract but article in French.
3. De Witt and Fortune 2019. Relationship-centred dementia care: insights from a community-based culture change coalition. *Reason for exclusion:* Focus not relationship based care or the Senses Framework.
4. Garcia-Castro et al. 2021. Life satisfaction and the mediating role of character strengths and gains in informal caregivers. *Reason for exclusion*: Focused on informal caregivers defined as an unpaid person (friend or relative) so the research is not in a care home setting.
5. Macaden et al. 2021. Relationship-centred CogniCare: an academic-digital-dementia care experts interface. *Reason for exclusion:* Not a care home or nursing home setting.
6. Metcalfe et al. 2012. Making sense of relationships. *Reason for exclusion:* Both men lived at home with their wives.
7. Milne 2009. Involving families in care homes: a relationship centred approach to dementia care. *Reason for exclusion:* Book review.
8. Morhardt 2013. From person-centred care to relational centred care. *Reason for exclusion:* Not based in care home or nursing home.
9. Morrison et al. 2019. Beyond tube feeding: relationship centred care, comfort care for individuals with eating challenges in dementia. *Reason for exclusion:* Letter to the editor.
10. Mullins et al: 2011. Experiences of spouses of people with dementia in long term care. *Reason for exclusion:* The focus is not on relationship based care or the senses framework.
11. Murphy et al. 2006. Research focus: relationship centred work with couples. *Reason for exclusion:* Not a care home environment.
12. Murphy 2010. Goodbye to the family from hell. *Reason for exclusion:* Course evaluation which discussed 2 themes. Not a nursing or care home setting.
13. Newcombe 2009. Involving families in care homes: a relationship centred approach to dementia care. *Reason for exclusion:* Book review.
14. Nwadiugwu et al. 2021. Early-onset dementia: key issues using a relationship-centred care approach. *Reason for exclusion:* This review focuses on early onset dementia and isn’t based within a care home.
15. Scerri et al. 2015. Discovering what works well: exploring quality dementia care in the hospital wards using an appreciative inquiry approach. *Reason for exclusion:* Not a care home or nursing home setting.
16. Song and Wei 2023. Doing relationship-centred dementia care: Learning from each other for better dementia support. *Reason for exclusion:* Book review.
17. Stewart et al. 2022. The senses framework: a relationship centred care approach to coproducing dementia events in order to allow people to live well after dementia diagnosis. *Reason for exclusion:* Not a care home or nursing home setting.
18. Tolman et al. 2018. A relationship-centred approach to managing pain in dementia. *Reason for exclusion:* Managing pain at home.

